# INCIDENCE AND FACTORS ASSOCIATED WITH LATE SPUTUM CULTURE CONVERSION AMONG MULTI- DRUG-RESISTANT TUBERCULOSIS PATIENTS ON TREATMENT IN NATIONAL REFERAL HOSPITAL UGANDA

**DOI:** 10.1101/2024.11.13.24317224

**Authors:** Cherop Adolphus, Nakiyingi Lydia, Joan Kalyango, Achiles Katamba, Ezekiel Mupere, Ssendikwanawa Emmanuel, Joan Rokani Bayowa, Cwinyaai Norman, Amutuhaire Judith Ssemasaazi, Okello Tom, Bagoloire Kolosi Lynn, Asilaza Vincent Kinya, Worodria William

**Affiliations:** Department of Clinical epidemiology, Makerere University College of Health Sciences, P.O. Box 7072, Kampala, Uganda; Clinical epidemiology and biostatistics department, College of Health Sciences, Makerere University, P.O. Box 7072, Kampala, Uganda; Epidemiology and Biostatistics Department, College of Health Sciences, Makerere University, P.O. Box 7072, Kampala, Uganda; School of Public Health, College of Health Sciences, Makerere university; Makerere Lung Institute, College of Health Sciences, Makerere University, Mulago National Referral Hospital; Infectious Disease Institute, Makerere lung institute, College of Health Sciences, Makerere University

## Abstract

**Background:** Tuberculosis (TB) remains one of the most common causes of death from an infectious disease. Late sputum culture conversion among Multi-Drug-Resistant Tuberculosis (MDR-TB) patients poses a risk for poor treatment outcomes. The study aimed to determine the incidence and factors associated with late sputum culture conversion among MDR-TB patients on treatment at the tuberculosis (TB) unit of Mulago National Referral Hospital.

**Methods:** A retrospective cohort study of 255 MDR-TB patient records between January 1^st^ 2012 to December 31^st^ 2018 were reviewed. Consecutive sampling was employed. Demographic characteristics, clinical factors and social factors were studied. STATA version 15 was used for analysis. Incidence was calculated as the ratio of MDR -TB patients with late sputum culture conversion result to the total number of participants studied. Factors associated were evaluated using generalized linear model (GLM) with Poisson family and log link using robust standard errors to adjust for over inflated variances.

**Results:** The incidence of late sputum culture conversion was 32% (95% CI 26.3-37.8). increasing age in years (incidence rate ratio IRR 1.004, 95%CI 1.000 1.008, P value 0.044), increasing weight (IRR 0.995, 95% CI 0.991-0.999, P value 0.020) and 9-12 months multi drug tuberculosis MDR-TB regimen (IRR 0.893, 95% CI 0.805-0.989, P value 0.030) were factors associated with late sputum culture conversion.

**Conclusions:** The incidence of late sputum culture conversion among MDR-TB patients on treatment was high (32%), occurring in about three out of every ten patients. Increasing weight, increasing age and 9-12 months MDR-TB drug regimen were significantly associated. Isolation of patients for a minimum of two months to minimize community transmission, starting eligible patients on the 9-12months MDR-TB regimen and categorization of patients into high-risk groups (elderly and underweight) with special targeted packages.

## Background

Tuberculosis (TB) remains one of the most common causes of death from an infectious disease worldwide (Murray et al., 2014). In 2019, an estimated 3.3% of new TB cases and 18% of previously treated cases had MDR/RR-TB equivalent to an estimated 465 000 (range, 400 000– 535 000) incident cases of rifampicin resistant TB; 78% had multidrug resistant TB (WHO, 2020).

The regional MDR-TB prevalence showed that Eastern Africa had an estimated burden of 1.7%; 1.1% – 2.2% cases (Musa et al., 2017) with drug resistant tuberculosis in Africa being largely missed with 93 000 cases estimated in 2016, only 27 828 (30%) were diagnosed (WHO, 2017).

Uganda remains among the high TB/HIV burden countries in the world. Multidrug resistant TB is an emerging problem with more than 1,040 estimated cases every year and the actual case finding is around 200 cases per year (MOH, 2017c). Incidence of Rifampicin Resistance/Multi Drug Resistant Tuberculosis (RR/MDR-TB) in 2017 was 4.7/100,000 population, translating to 1900 incident RR/MDR-TB cases and the country notified 489 incident RR/MDR-TB cases, which was only 26% (489/1,900) of the estimated cases (MOH, 2017a). Among this 1.6% (0.87–2.5) were new cases, 12% (6.5–19) were previously treated (MOH,2017b). Knowing the time to sputum culture conversion is often used as an early predictor for treatment outcomes especially in MDR-TB patients, and a delay in conversion greater than one month was considered as a suspect of MDR-TB treatment failure (Yihunie Akalu et al., 2018) (Kurbatova et al., 2012), (Mulu et al., 2015) (Velayutham et al., 2016). Resistance to anti-tuberculosis drugs, causes worse treatment outcomes, infectiousness, failure and XDR TB) and mortality (Kurbatova et al., 2015, WHO, 2018) (EVANGELISTA et al., 2018). Monitoring the outcomes of TB treatment involves obtaining sputum for microscopic examination and sputum culture at monthly intervals (MOH, 2017). Uganda national tuberculosis and leprosy policy (NTLP) recommends that drug sensitivity tests (DST) and sputum culture should be performed monthly before smear and sputum culture conversion. Different time to sputum culture conversion and factors influencing sputum culture conversion have been reported, most commonly are diabetes mellitus (Magee et al., 2014a), body mass index (BMI) (Putri et al., 2014b), sputum smear grade before the treatment (Unsematham and Kateruttanakul, 2013), vitamin D concentration (Arnedo-Pena et al., 2015) and various social factors (smoking and alcoholism) (Arnedo-Pena et al., 2015), MDR-TB category, HIV co-infection, presence of radiological findings. There are controversies in studies conducted as different time to sputum culture conversion and associated factors have been reported for instance studies by (Filate et al., 2018)and (Gunda et al., 2017) found that male gender and age >50 years were associated and (Putri et al., 2014b) found female sex as a predictor for longer sputum culture conversion while (Akalu et al., 2018) found that the median sputum culture conversion time in this study was 65 (60-70 days). (Qazi et al., 2011) found that the median time to culture conversion was 196 days (range 32–471). However, there is limited data on MDR-TB culture conversion in our setting and associated factors. With several controversial reports from other settings and the scanty of data in our setting, we sought to determine the incidence and associated factors with late sputum culture conversion among MDR-TB patients in Uganda. Therefore, given the inconsistencies reported in other studies and scanty data in our setting about late sputum culture conversion among MDR-TB patients in Uganda, the study aimed to determine the incidence and factors associated with late sputum culture conversion among multi drug resistant tuberculosis patients on treatment in Uganda.

## Methods

### Design and setting

This was a retrospective cohort study of MDR-TB patients attending Mulago National Referral Hospital MDR-TB treatment unit.

### Study participants

A sample of 255 MDR-TB patients records with a positive baseline sputum culture result were consecutively sampled from January 1^st^, 2012, to December 31^st^, 2018. Data was accessed from 10^th^ May 2021to 1^st^june 2021 and no authors had access to information that could identify individual participants during or after data collection. On average the clinic receives an estimated 115 patients annually with a total of 693 from 2012 to 2018.

### Sample size calculation

Consecutive sampling was employed. The sample size was estimated using Kish Leslie formula for objective one and sample size for comparison of two proportions for objective two. The overall sample size was 255 patient records.

### Study procedures

An abstraction form composed of study variables was used to collect data from patient registers and files by the principal investigator and three well-trained research assistants. Data was collected from MDR-TB registers and patient files. Twenty patients’ records were used for pre-testing and independent variables included demographic factors (age, sex, and area of residence), clinical factors {body weight, category of patient (new TB case, TB Re-treatment case, TB relapse, TB defaulter), drug regimen, re-treatment, bacteria Load, adherence, side effects and HIV/AIDS} and social factors (smoking and alcoholism). Dependent variable was sputum culture conversion greater than 2 months. MDR-TB patient who had a baseline positive sputum culture at the start of MDR-TB treatment but remained positive at greater than 2 months. Data on the above variables was collected by the principal investigator and the research assistants.

### Definition of variables

Sputum culture conversion: Defined as two negative consecutive cultures taken at least 30 days apart. Late conversion: MDR-TB patient who had a baseline positive sputum culture at the start of MDR-TB treatment but remained positive at more than 2 months.

### Data analysis

A database was set up using Epi data version 3.2 and data was double entered by principal investigator and one of the research assistants, then exported to STATA version 15 for analysis.

#### Descriptive analysis

participant’s demographic factors were described using percentages for categorical variables, median and interquartile ranges for skewed distribution and mean and standard deviation for normal distribution. Incidence of late sputum culture conversion among multi drug resistant tuberculosis patients was computed as the ratio of multi drug resistant confirmed tuberculosis patients with a late negative sputum culture results to the total number of participants followed up. late sputum culture conversion was sputum conversion greater than 2 months. Factors associated with late sputum conversion were evaluated using generalized linear model with Poisson family and log link and using robust standard errors to adjust for over inflated variances.

## Ethics consideration

Permission to the study and Letter for waiver of consent to use the tuberculosis unit register was granted by Makerere University Clinical Epidemiology Unit. Ethical approval from School of Medicine Research and Ethics Committee (SOMREC) and Mulago national referral hospital administrative approval.

## Results

### Socio-demographic characteristics

A total of 255 patients were studied and majority of the respondents were male (62.4%), weight was normally distributed with a mean of 51.3 (SD = 9.6) kilograms and the median age was 30 years (IQR = 24 - 39). Less than half (43.1) % were married (table 1). Majority of participants had good adherence (80%, n= 178) and were on a 6-24 regimen (79.6% n=203). About 32.9% (n=84) were alcohol consumers and 1.6% (n=4) had comorbidities (cancer and diabetes) and 54.9% were HIV/AIDS positive.

**Table 1.**
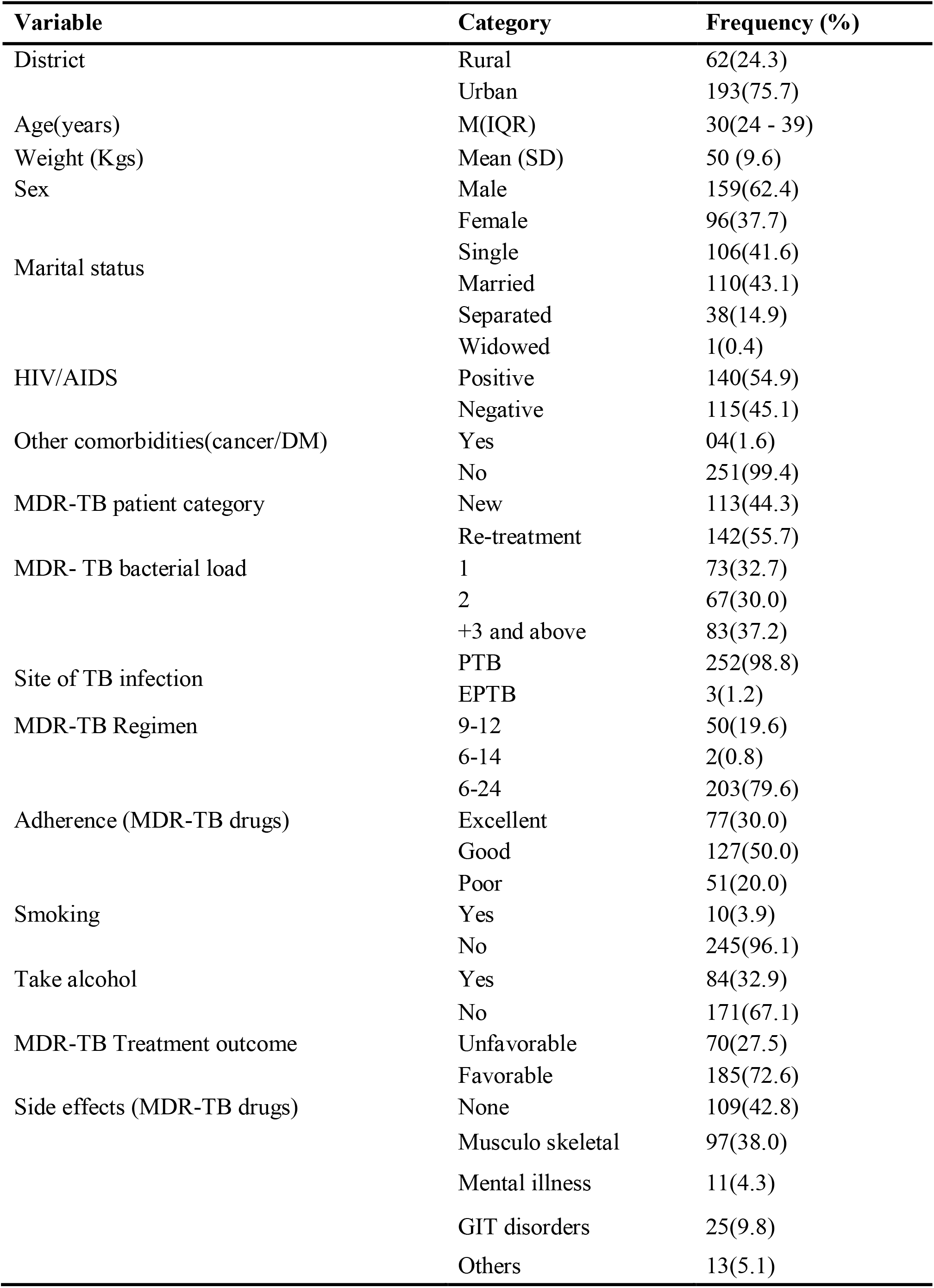
study participant characteristic.

**Table 2.** Incidence of sputum culture conversion among 255 participants at Mulago tuberculosis unit from 2012 to 2018.

| Overall period (months) | Percentage (%) | 95% CI |
| --- | --- | --- |
| 1-2 | 68.2 | 62.2 -73.7 |
| >2 | 31.8 | 25.3- 40.3 |

**Table 3.** Multivariate analysis for factors associated with late sputum culture conversion.

| Variable | Category | IRR | 95% | P value |
| --- | --- | --- | --- | --- |
| Age | N/A | 1.004 | 1.000 1.010 | 0.042 |
| Weight | N/A | 0.995 | 0.990 0.998 | 0.004 |
| Regimen | 6-24 | 1 |  |  |
|  | 9-12 | 0.893 | 0.806 0.990 | 0.032 |

Majority of the patients were re-treatments (55.7%), had a bacterial load of +3 and site of infection was pulmonary tuberculosis.

### Incidence

Late sputum culture converters were MDR-TB patients who converted sputum culture at more than 2 months of treatment. The overall incidence of late sputum conversion among MDR-TB patients in care and treatment from the period of 2012 to 2018 was 32% (81/255) (95% CI 26.3-37.8%).

## Discussion

This research examined the incidence and factors associated with late sputum culture conversion among MDR-TB patients on treatment in Mulago National Referral Hospital MDR-TB unit. In this study the incidence rate of late sputum culture conversion was 32% (95% CI 26.7-38.2). Age weight and MDR-TB regimen independently predicted late sputum culture conversion among multi-drug-resistant tuberculosis patients on treatment in Uganda.

### Late sputum conversion

The study revealed that the incidence of late sputum culture conversion among multi drug resistant tuberculosis patients on treatment was high occurring in about three in every ten patients 32% 95% CI 26.7-38.2. This incidence was higher than the national incidence of 0.161% (161/100,000) (Jawairia et al., 2017). Studies done by Liu et al, 2018 in China, Basit et al, 2014 in Pakistan, Javid et al, 2018 in Peshawar and Tekalegn et al, 2017 in Ethiopia showed a higher incidence of late sputum culture conversion 76.7%, 45,7%, 35.9% and 47% respectively. This difference for China can be explained by the fact the studies were conducted in different cities with high burden of tuberculosis and all were urban centers. However, in our study the introduction of Regional Referrals as treatment centers might have contributed to the reduction in the volume of patients. China being highly populated and developed than Uganda which means the population is at high risk of MDR-TB spread which explains the higher incidence. For Pakistan, the study was conducted in the only MDR-TB treatment site in four provinces which implies that the burden was high and so the high incidence. For Javid et al in Peshawar there is a high burden with retreatments contributing about 18.1% and new 3.7% in a survey hence the high incidence of late conversion. A study by <u>Shibabaw</u> in Ethiopia revealed that 35.9% of MDR-TB patients late converted sputum culture which was similar to our finding and this can be explained by the similarity in sample size and study setting as both were Regional Referrals and in urban areas with almost the same TB burden of 192 per 100,000 and 200 per 100,000 respectively. However, a study by Assemie et al., 2020 conducted in East Africa found a lower incidence which can be explained by the fact that the study design was a review of literature and the differences in the distribution of the MDR-TB within the East African region which is composed of Ethiopia, Kenya, Mozambique, Tanzania, Zambia, Zimbabwe, and Somalia. Some regions may have a very low incidence and so results in the lower incidence.

Delaying sputum culture conversion results in economic wastage by prolonging the duration of treatment, poor treatment adherence and consequently failing treatment. It is also associated with higher case fatality rates (50–80%) as a result of drug toxicity, leading to the emergence of XDR-TB (Kurbatova et al., 2012) (Kim et al., 2016). This exposes the community at risk of cross infection.

### Associated factors

#### Increasing weight

The weight in kilograms of patients as a measure of determining the dosage of the multi drug resistance tuberculosis treatment is key in management and was found to be significantly associated with late sputum conversion (RR 0.994 95% CI 0.991 0.999 p-value 0.020). For every unit increase in weight, there was 1% decrease in the likelihood of having late sputum culture conversion. This can be explained by the fact that weight is core in determining the dosage of treatment using milligrams per kilogram body weight. So, with increasing weight, dosage increases, and this increases the pressure on the mycobacterium tuberculosis hence less likelihood of late sputum culture conversion. This was in agreement with studies by Putri, Assemie (China), Park (South Korea)and Magee (Georgia) that revealed that severe underweight (BMI <16 kg/m(2) was associated with longer time to initial sputum culture conversion among MDR-TB patients (Putri et al., 2014a) (Assemie et al., 2020) (Park et al., 2016) (Magee et al., 2014b). A low BMI directly relates to low body weight.

#### Increasing age in years

Age was associated with late sputum culture conversion (RR 1.004 95CI 1.00-1.009 p-value 0.044). For every unit increase in age, there was a 0.4 more likelihood of having a late sputum culture conversion. This implies that as one grows older, the higher the chances of late sputum culture conversion. This was similar to study findings in Tanzania by Daniel Gunda which found age > 50 years to be associated OR 6.7 p-value 0.003 (Gunda et al., 2017). This can be explained by the fact that as a person grows older, the body immunity lowers hence susceptible to late sputum culture conversion. However, a study by Liu in china found no association with age which was inconsistent with our study findings.

#### MDR-TB regimen

MDR-TB Regimen 9-12 months was also associated with late sputum culture conversion (RR = 0.893 95% CI 0.805 0.999 p-value = 0.030). Patients on 9-14 months MDR-TB regimen had a 11% reduced likelihood of late sputum culture conversion. This result was consistent with a multi-center study which revealed that receiving an average of at least six potentially effective drugs was associated with a 36% greater likelihood of sputum culture conversion than receiving an average of at least five but fewer than six potentially effective drugs per day (Kurbatova et al., 2015). This can be explained by the fact that 9-14 based regimen is a short duration regimen with new drugs like bedaqulline which patients may have never had exposure to and so more effective as it gives much more pressure on the bacteria which increased the chances of converting sputum.

## Conclusions

The occurrence of late sputum culture conversion reported as incidence was high at about three in every ten patients. Increasing age, increasing weight in kilograms and 9-12 months MDR-TB regimen were independent factors of late sputum culture conversion among multi drug resistant tuberculosis patients on treatment in Uganda.

## Data Availability

N/A

## Abbreviations

AIDS: Acquired Immune Deficiency Syndrome
BMI: Body Mass Index
CI: Confidence Interval
DST: Drug Susceptibility Test
EPTB: Extra Pulmonary Tuberculosis
FLD: First Line Drugs
HIV: Human Immuno deficiency Virus
HR: Hazard Ratio
IRR: Incidence Rate Ratio
LJ: Lowenstein Jensen
MDR: Multi Drug Resistance
MGIT: Mycobacterium Growth Indicator Tube
MOH: Ministry of Health
NTLP: National Tuberculosis and Leprosy Programme
OR: Odds Ratio
PTB: Pulmonary Tuberculosis
RR: Rifampicin Resistance
SCC: Sputum Culture Conversion
SOMREC: School of Medicine Research and Ethics Committee
TB: Tuberculosis
USAID: United States Agency for International Development
WHO: World Health Organization
XDR: Extensively Drug Resistant.

## Acknowledgements

The author would like to acknowledge the invaluable contribution made bythe supervisors Dr. Nakinyingi Lydia and Dr. Worodria William, research assistants and Mulago hospital staff. Special thanks to Clinical epidemiology unit staff for the technical support and MAPRONANO ACE for the financial support.

## Participant flow chart

**Fig 1.**
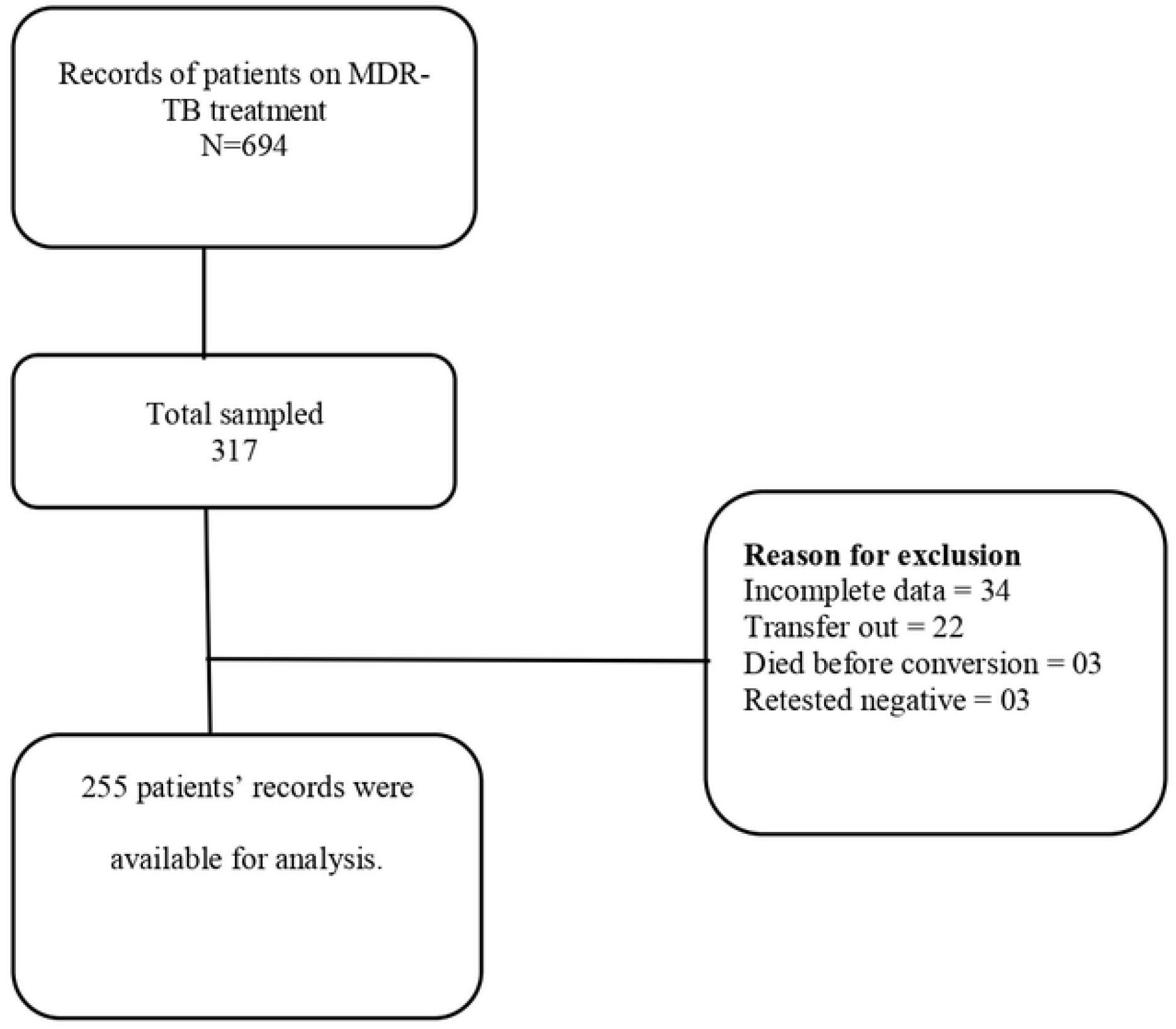
Participant flow diagram.

## References

Akalu, Y., Temesgen, M., Kindie, F. & Alemu Gelaye, K. 2018. Time to sputum culture conversion and its determinants among Multi-drug resistant Tuberculosis patients at public hospitals of the Amhara Regional State: A multicenter retrospective follow up study. PloS one, 13, e0199320–e0199320.

Arnedo-Pena, A., Juan-Cerdan, J. V., Romeu-Garcia, M. A., Garcia-Ferrer, D., Holguin-Gomez, R., Iborra-Millet, J. & Pardo-Serrano, F. 2015. Vitamin D status and incidence of tuberculosis infection conversion in contacts of pulmonary tuberculosis patients: a prospective cohort study. Epidemiol Infect, 143, 1731–41.

Assemie, M. A., Alene, M., Petrucka, P., Leshargie, C. T. & Ketema, D. B. 2020. Time to sputum culture conversion and its associated factors among multidrug-resistant tuberculosis patients in Eastern Africa: A systematic review and meta-analysis. International Journal of Infectious Diseases, 98, 230–236.

DO SOCORRO NANTUA EVANGELISTA, M., Maia, R., Toledo, J. P., De Abreu, R. G., Braga, J. U., Barreira, D. & Trajman, A. 2018. Second month sputum smear as a predictor of tuberculosis treatment outcomes in Brazil. BMC research notes, 11, 414–414.

Evangelista, D. S. N. M., Maia, R., Toledo, J. P., De Abreu, R. G., Braga, J. U., Barreira, D. & Trajman, A. 2018. Second month sputum smear as a predictor of tuberculosis treatment outcomes in Brazil. BMC hematology, 11, 414–414.

Filate, M., Mehari, Z. & Alemu, Y. M. 2018. Longitudinal body weight and sputum conversion in patients with tuberculosis, Southwest Ethiopia: a retrospective follow-up study. BMJ Open, 8, e019076.

Gunda, D. W., Nkandala, I., Kavishe, G. A., Kilonzo, S. B., Kabangila, R. & Mpondo, B. C. 2017. Prevalence and Risk Factors of Delayed Sputum Conversion among Patients Treated for Smear Positive PTB in Northwestern Rural Tanzania: A Retrospective Cohort Study. Journal of Tropical Medicine, 2017, 5352906.

Kim, J., Kwak, N., Lee, H. Y., Kim, T. S., Kim, C.-K., Han, S. K. & Yim, J.-J. 2016. Effect of drug resistance on negative conversion of sputum culture in patients with pulmonary tuberculosis. International Journal of Infectious Diseases, 42, 64–68.

Kurbatova, E. V., Cegielski, J. P., Lienhardt, C., Akksilp, R., Bayona, J., Becerra, M. C., Caoili, J., Contreras, C., Dalton, T., Danilovits, M., Demikhova, O. V., Ershova, J., Gammino, V. M., Gelmanova, I., Heilig, C. M., Jou, R., Kazennyy, B., Keshavjee, S., Kim, H. J., Kliiman, K., Kvasnovsky, C., Leimane, V., Mitnick, C. D., Quelapio, I., Riekstina, V., Smith, S. E., Tupasi, T., Van Der Walt, M., Vasilyeva, I. A., Via, L. E., Viiklepp, P., Volchenkov, G., Walker, A. T., Wolfgang, M., Yagui, M. & Zignol, M. 2015. Sputum culture conversion as a prognostic marker for end-of-treatment outcome in patients with multidrug-resistant tuberculosis: a secondary analysis of data from two observational cohort studies. The Lancet Respiratory Medicine, 3, 201–209.

Kurbatova, E. V., Gammino, V. M., Bayona, J., Becerra, M. C., Danilovitz, M., Falzon, D., Gelmanova, I., Keshavjee, S., Leimane, V., Mitnick, C. D., Quelapio, M. I., Riekstina, V., Taylor, A., Viiklepp, P., Zignol, M. & Cegielski, J. P. 2012. Predictors of sputum culture conversion among patients treated for multidrug-resistant tuberculosis. Int J Tuberc Lung Dis, 16, 1335–43.

Magee, M. J., Kempker, R. R., Kipiani, M., Tukvadze, N., Howards, P. P., Narayan, K. M. & Blumberg, H. M. 2014a. Diabetes mellitus, smoking status, and rate of sputum culture conversion in patients with multidrug-resistant tuberculosis: a cohort study from the country of Georgia. PLoS One, 9, e94890.

Magee, M. J., Kempker, R. R., Kipiani, M., Tukvadze, N., Howards, P. P., Narayan, K. M. V. & Blumberg, H. M. 2014b. Diabetes mellitus, smoking status, and rate of sputum culture conversion in patients with multidrug-resistant tuberculosis: a cohort study from the country of Georgia. PloS one, 9, e94890–e94890.

MOH 2017a. National TB Conference 2018: Stakeholders call for increased domestic funding towards Tuberculosis.

MOH, N. T. A. L. P. N. 2017b. Uganda Tuberculosis profile.

MOH, U. N. T. A. L. C. P. 2017c.

MANUAL FOR MANAGEMENT and CONTROL OF TUBERCULOSIS and LEPROSY 3RD EDITION MARCH 2017.

Mulu, W., Mekonnen, D., Yimer, M., Admassu, A. & Abera, B. 2015. Risk factors for multidrug resistant tuberculosis patients in Amhara National Regional State. Afr Health Sci, 15, 368–77.

Murray, C. J., Ortblad, K. F., Guinovart, C., Lim, S. S., Wolock, T. M., Roberts, D. A., Dansereau, E. A., Graetz, N., Barber, R. M., Brown, J. C., Wang, H., Duber, H. C., Naghavi, M., Dicker, D., Dandona, L., Salomon, J. A., Heuton, K. R., Foreman, K., Phillips, D. E., Fleming, T. D., Flaxman, A. D., Phillips, B. K., Johnson, E. K., Coggeshall, M. S., Abd-Allah, F., Abera, S. F., Abraham, J. P., Abubakar, I., Abu-Raddad, L. J., Abu-Rmeileh, N. M., Achoki, T., Adeyemo, A. O., Adou, A. K., Adsuar, J. C., Agardh, E. E., Akena, D., Al Kahbouri, M. J., Alasfoor, D., Albittar, M. I., Alcala-Cerra, G., Alegretti, M. A., Alemu, Z. A., Alfonso-Cristancho, R., Alhabib, S., Ali, R., Alla, F., Allen, P. J., Alsharif, U., Alvarez, E., Alvis-Guzman, N., Amankwaa, A. A., Amare, A. T., Amini, H., Ammar, W., Anderson, B. O., Antonio, C. A., Anwari, P., Arnlov, J., Arsenijevic, V. S., Artaman, A., Asghar, R. J., Assadi, R., Atkins, L. S., Badawi, A., Balakrishnan, K., Banerjee, A., Basu, S., Beardsley, J., Bekele, T., Bell, M. L., Bernabe, E., Beyene, T. J., Bhala, N., Bhalla, A., Bhutta, Z. A., Abdulhak, A. B., Binagwaho, A., Blore, J. D., Basara, B. B., Bose, D., Brainin, M., Breitborde, N., Castaneda-Orjuela, C. A., Catala-Lopez, F., Chadha, V. K., Chang, J. C., Chiang, P. P., Chuang, T. W., Colomar, M., Cooper, L. T., Cooper, C., Courville, K. J., Cowie, B. C., Criqui, M. H., Dandona, R., Dayama, A., De Leo, D., Degenhardt, L., Del Pozo-Cruz, B., Deribe, K., et al. 2014. Global, regional, and national incidence and mortality for HIV, tuberculosis, and malaria during 1990-2013: a systematic analysis for the Global Burden of Disease Study 2013. Lancet, 384, 1005–70.

Musa, B. M., Adamu, A. L., Galadanci, N. A., Zubayr, B., Odoh, C. N. & Aliyu, M. H. 2017. Trends in prevalence of multi drug resistant tuberculosis in sub-Saharan Africa: A systematic review and meta-analysis. PLoS One, 12, e0185105.

Putri, F., Burhan, E., Nawas, A., Soepandi, P., Kusumo Sutoyo, D., Agustin, H., Isbaniah, F. & Dowdy, D. 2014a. Body mass index predictive of sputum culture conversion among MDR-TB patients in Indonesia. The International Journal of Tuberculosis and Lung Disease, 18, 564–70.

Putri, F. A., Burhan, E., Nawas, A., Soepandi, P. Z., Sutoyo, D. K., Agustin, H., Isbaniah, F. & Dowdy, D. W. 2014b. Body mass index predictive of sputum culture conversion among MDR-TB patients in Indonesia. Int J Tuberc Lung Dis, 18, 564–70.

Qazi, F., Khan, U., Khowaja, S., Javaid, M., Ahmed, A., Salahuddin, N., Hussain, H., Becerra, M. C., Golub, J. E. & Khan, A. J. 2011. Predictors of delayed culture conversion in patients treated for multidrug-resistant tuberculosis in Pakistan. The international journal of tuberculosis and lung disease : the official journal of the International Union against Tuberculosis and Lung Disease, 15, 1556–i.

Unsematham, S. & Kateruttanakul, P. 2013. Factors predicting sputum smear conversion and treatment outcomes in new smear-positive pulmonary tuberculosis. J Med Assoc Thai, 96, 644–9.

Velayutham, B., Nair, D., Kannan, T., Padmapriyadarsini, C., Sachdeva, K. S., Bency, J., Klinton, J. S., Haldar, S., Khanna, A., Jayasankar, S. & Swaminathan, S. 2016. Factors associated with sputum culture conversion in multidrug-resistant pulmonary tuberculosis. Int J Tuberc Lung Dis, 20, 1671–1676.

WHO 2017. WHO Global TB report. 2017.

WHO 2018. Antimicrobial Resistance. Geneva: World Health Organization

WHO 2020. Global tuberculosis report world health organisation.

Yihunie Akalu, T., Muchie, K. F. & Alemu Gelaye, K. 2018. Time to sputum culture conversion and its determinants among Multi-drug resistant Tuberculosis patients at public hospitals of the Amhara Regional State: A multicenter retrospective follow up study. PloS one, 13, e0199320–e0199320.

